# Seasonal human coronavirus antibodies are boosted upon SARS-CoV-2 infection but not associated with protection

**DOI:** 10.1101/2020.11.06.20227215

**Authors:** Elizabeth M. Anderson, Eileen C. Goodwin, Anurag Verma, Claudia P. Arevalo, Marcus J. Bolton, Madison E. Weirick, Sigrid Gouma, Christopher M. McAllister, Shannon R. Christensen, JoEllen Weaver, Phillip Hicks, Tomaz B. Manzoni, Oluwatosin Oniyide, Holly Ramage, Divij Mathew, Amy E. Baxter, Derek A. Oldridge, Allison R. Greenplate, Jennifer E. Wu, Cécile Alanio, Kurt D’Andrea, Oliva Kuthuru, Jeanette Dougherty, Ajinkya Pattekar, Justin Kim, Nicholas Han, Sokratis A. Apostolidis, Alex C. Huang, Laura A. Vella, The UPenn COVID Processing Unit, E. John Wherry, Nuala J. Meyer, Sara Cherry, Paul Bates, Daniel J. Rader, Scott E. Hensley

## Abstract

Severe acute respiratory syndrome coronavirus 2 (SARS-CoV-2) has rapidly spread within the human population. Although SARS-CoV-2 is a novel coronavirus, most humans had been previously exposed to other antigenically distinct common seasonal human coronaviruses (hCoVs) before the COVID-19 pandemic. Here, we quantified levels of SARS-CoV-2-reactive antibodies and hCoV-reactive antibodies in serum samples collected from 204 humans before the COVID-19 pandemic. We then quantified pre-pandemic antibody levels in serum from a separate cohort of 252 individuals who became PCR-confirmed infected with SARS-CoV-2. Finally, we longitudinally measured hCoV and SARS-CoV-2 antibodies in the serum of hospitalized COVID-19 patients. Our studies indicate that most individuals possessed hCoV-reactive antibodies before the COVID-19 pandemic. We determined that ∼23% of these individuals possessed non-neutralizing antibodies that cross-reacted with SARS-CoV-2 spike and nucleocapsid proteins. These antibodies were not associated with protection against SARS-CoV-2 infections or hospitalizations, but paradoxically these hCoV cross-reactive antibodies were boosted upon SARS-CoV-2 infection.

## INTRODUCTION

Coronaviruses commonly infect humans^1-4^. The severe acute respiratory syndrome coronavirus 2 (SARS-CoV-2) emerged at the end of 2019 and has rapidly spread among humans, many of whom have been previously exposed to common seasonal human coronaviruses (hCoVs)^5^. Common seasonal hCoVs include the betacoronaviruses HKU1 and OC43 and the alphacoronaviruses 229E and NL63^6-9^. SARS-CoV-2 belongs to the betacoronavirus genus and is more closely related to HKU1 and OC43 compared to the alphacoronaviruses 229E and NL63^10^. A recent study examining electronic medical records concluded that recent hCoV infections are not associated with decreased SARS-CoV-2 infections, but are associated with reducing the severity of Coronavirus Disease 2019 (COVID-19)^11^. It is unknown if prior hCoV exposures elicit antibodies that prevent or alter the outcomes of SARS-CoV-2 infections. Further, it is unknown if different aged individuals have distinct hCoV immune histories that can affect SARS-CoV-2 susceptibility. To address this, we completed a serological survey using serum samples collected from different aged humans prior to the COVID-19 pandemic. We quantified levels of antibodies reactive to viral proteins from hCoVs and determined if these antibodies were associated with SARS-CoV-2 protection. Finally, we completed a series of studies using serum collected from COVID-19 patients to determine if antibodies reactive to hCoVs are boosted upon SARS-CoV-2 infections.

## RESULTS

### Identification of SARS-CoV-2-reactive Antibodies in Human Sera Collected Prior to the COVID-19 Pandemic

We completed ELISAs to quantify levels of pre-pandemic SARS-CoV-2-reactive IgG antibodies in 204 human serum samples collected in 2017. We tested serum samples collected from 36 children (age 1-17) at the Children’s Hospital of Philadelphia originally collected for lead testing and 168 adults (age 18-90) who had been recruited into the Penn Medicine Biobank. We tested Penn Medicine Biobank samples from individuals who had no medical history of cancer or organ transplantation, pregnancy during the previous 9 months, or an infectious disease within the previous 28 days prior to blood draw. Using these samples, we previously found that different aged individuals possess H3N2 influenza virus antibodies that have different specificities^12^.

We found that 5.4% of serum samples collected in 2017 contained IgG antibodies that reacted to the SARS-CoV-2 full length spike (S) protein (**Figure 1a**), 2.0% of samples contained antibodies that reacted to the receptor binding domain (RBD) of the SARS-CoV-2 S protein (**Figure 1b**), and 18.6% of samples contained antibodies that reacted to the SARS-CoV-2 nucleocapsid (N) protein (**Figure 1c**). Several pre-pandemic serum samples contained antibodies that were at similar levels as those in serum from PCR-confirmed COVID-19 recovered donors (**Figure 1a-c**). Most serum samples with antibodies reactive to the SARS-CoV-2 full length S protein did not have antibodies that reacted to the SARS-CoV-2 S-RBD protein (**Figure 1d**), which is consistent with a recent study showing that some individuals possessed pre-pandemic antibodies against the S2 domain of the SARS-CoV-2 S protein^13^. In contrast to serum antibodies isolated from PCR-confirmed COVID-19 recovered donors, serum antibodies from individuals collected before the pandemic had very low or undetectable levels of SARS-CoV-2 neutralizing antibodies, regardless of whether or not the sample possessed cross-reactive antibodies against SARS-CoV-2 S and N proteins (**Figure 1e**). We found no obvious differences in levels of SARS-CoV-2 cross-reactive antibodies among donors with different birth years (**Figure S1 a-c**).

**Figure 1.**
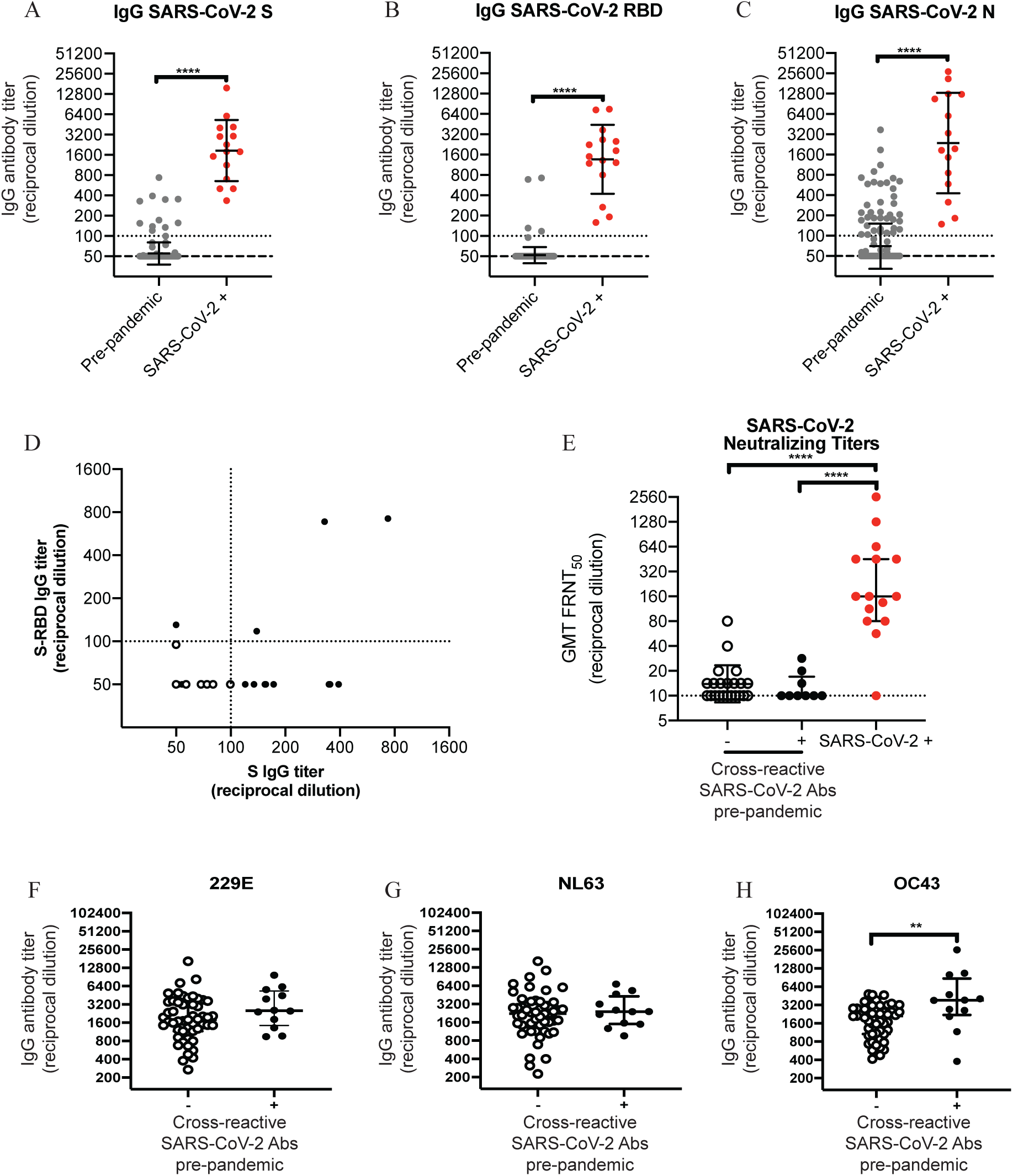
Identification of pre-existing cross-reactive SARS-CoV-2 antibodies in human serum prior to the pandemic. ELISAs were completed to quantify levels of serum antibodies binding to the SARS-CoV-2 full-length spike (S) protein (**A**), the receptor binding domain (S-RBD) of S (**B**), and the nucleocapsid (N) protein (**C**); dashed line denotes lower limit of detection (LOD=50), dotted line represents a threshold set 2-fold above LOD (>100). We tested samples collected from 204 individuals in the summer of 2017, prior to the global pandemic. We also tested samples collected from 15 individuals following confirmed SARS-CoV-2 infections. and recovered adults. (**D**) The relationship between antibody titers in donors with detectable IgG against the S-RBD and/or full length S is shown. (**E**) SARS-CoV-2 pseudotype neutralization assays were completed using pre-pandemic serum samples with (n=9) and without (n=22) cross reactive SARS-CoV-2 antibodies, as well as serum samples from individuals following confirmed SARS-CoV-2 infections (n=15); one-way ANOVA Tukey’s multiple comparisons of log2 transformed antibody titers ****p<0.0001; dotted line denotes lower LOD (=10). (**F**-**H**) ELISAs were completed to quantify levels of serum antibodies binding to the full length S proteins from 229E, NL63, and OC43 using pre-pandemic serum samples with (n=12) and without (n=51). Unpaired t-tests of log2 transformed antibody titers ****p<0.0001 and **p=0.0027. Horizontal lines indicate geometric mean and error bars represent standard deviation.

### Humans with Pre-pandemic SARS-CoV-2-reactive Antibodies Had Elevated Levels of Antibodies Against Previously Circulating Betacoronaviruses

We completed ELISAs to quantify levels of pre-pandemic hCoV-reactive IgG antibodies in all 204 human serum samples collected in 2017. Most serum samples possessed antibodies that reacted to the S protein of 229E and NL63 (both alphacoronaviruses), as well as OC43 (a betacoronavirus) (**Figure S1d-f**). There were no major differences in levels of these antibodies among individuals with different birth years, however serum from very young children possessed lower levels of antibodies reactive to the 229E and NL63 S proteins (**Figure S1d-f**). We completed full antibody titrations to directly compared levels of hCoV antibodies in a subset of pre-pandemic samples from individuals who either did (n=12) or did not (n=51) possess cross-reactive SARS-CoV-2 antibodies (**Figure 1f-h**). Pre-pandemic antibody levels against the 229E and NL63 alphacoronavirus S proteins were similar among individuals with and without SARS-CoV-2 reactive antibodies (**Figure 1f-g**). In contrast, antibody levels against the betacoronavirus OC43 S protein were higher in individuals with SARS-CoV-2 reactive antibodies compared to individuals who did not possess pre-pandemic SARS-CoV-2 reactive antibodies (**Figure 1h**). These data suggest that pre-pandemic SARS-CoV-2 reactive antibodies were likely elicited by previously circulating betacoronavirus strains, such as OC43.

### Pre-existing hCoV Cross-reactive Antibodies Were Not Associated With Protection From SARS-CoV-2 Infections

It is unknown if antibodies elicited by prior hCoV infections protect against SARS-CoV-2 infections and/or prevent severe COVID-19. To address this, we measured SARS-CoV-2 IgG antibodies in pre-pandemic serum samples from 251 individuals who subsequently went on to become PCR-confirmed infected with SARS-CoV-2 and in a control group of pre-pandemic samples from 251 matched individuals who did not become infected with SARS-CoV-2. Pre-pandemic samples were collected by the Penn Medicine BioBank from August 2013 to March 2020 and PCR-confirmed SARS-CoV-2 infections were identified by nasopharyngeal swab PCR testing results in electronic health records. We found that 2.2% samples possessed pre-pandemic antibodies reactive to the SARS-CoV-2 full length S protein, 0.6% samples possessed pre-pandemic antibodies reactive to the SARS-CoV-2 S-RBD, and 23.9% samples possessed pre-pandemic antibodies reactive to the SARS-CoV-2 N protein. Importantly, we found no differences in SARS-CoV-2-reactive antibodies in serum samples from individuals who did or did not become subsequently infected with SARS-CoV-2 (**Figure 2a**; S protein: p=0.62, S-RBD: p=0.49, N protein: p=0.34 and **Table S1** and **Table S2**). We also measured antibodies reactive to the OC43 S protein and found no differences among samples from individuals who did or did not become infected with SARS-CoV-2 (**Figure 2a**; p=0.90 and **Table S1** and **Table S2**). Among those with PCR-confirmed SARS-CoV-2 infections, we found no relationship between antibody titers and hospitalization or disease severity among hospitalized patients (**Table S1** and **Table S2**). We found no relationship between antibody titers and the need for respiratory support and admittance into the ICU following SARS-CoV-2 infection (**Table S1** and **Table S2**).

**Figure 2.**
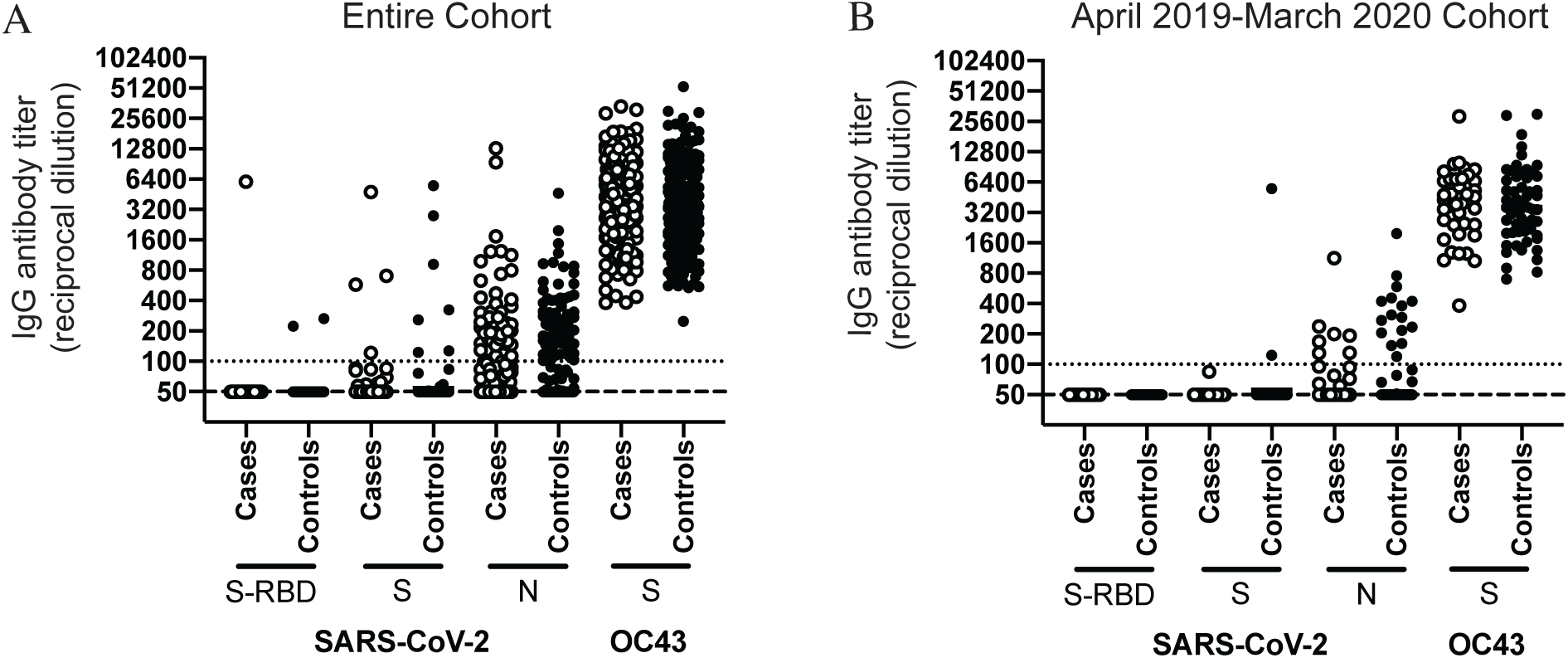
Pre-pandemic SARS-CoV-2 and OC43-reactive antibodies are not associated with protection from SARS-CoV-2 infection. We quantified antibody levels in pre-pandemic serum samples collected from individuals who later became SARS-CoV-2 infected (cases; n=251) and those who did not become SARS-CoV-2 infected (controls; n=251). ELISAs were completed to quantify levels of antibodies reactive to SARS-CoV-2 proteins (S, S-RBD, and N) and the OC43 S protein. Shown are data using samples collected from the entire cohort between August 2013 and March 2020 (**A**) and samples from a smaller subset of individuals collected between April 2019-Mach 2020 (**B**). Antibody titers between cases and controls were not significantly different as determined by unpaired t-tests of log2 transformed antibody titers. Dashed line denotes lower limit of detection (LOD=50), dotted line represents a threshold set 2-fold above LOD (>100).

Previous studies indicated that immunity to hCoV can be short-lived^14^ and a recent study documented that antibody titers against hCoV can fluctuate over time^5^, presumably due to repetitive hCoV exposures. In our study, pre-pandemic serum samples were collected from 2013-2020 and therefore it is possible that antibody levels in some of the samples collected several years prior to 2020 do not accurately reflect antibody levels present during the COVID-19 pandemic. To address this, we compared SARS-CoV-2 and OC43 IgG antibody titers in the serum of individuals in our cohort who had samples collected within one year of the pandemic (between April 2019 and March 2020). Using this smaller cohort (n=39 SARS-CoV-2 cases and n=57 controls), we still found no differences in levels of antibodies reactive to the SARS-CoV-2 S protein, S-RBD protein, N protein, or OC43 S protein (**Figure 2**B). Taken together, our data suggest that a subset of humans possessed non-neutralizing cross-reactive antibodies against SARS-CoV-2 S and N proteins prior to the COVID-19 pandemic, but these antibodies were not associated with protection from SARS-CoV-2 infections or reducing hospitalizations upon SARS-CoV-2 infections.

### SARS-CoV-2 Boosts Antibodies Reactive to Other Human Betacoronaviruses

Recent studies indicate that COVID-19 recovered donors possess higher levels of antibodies against seasonal betacoronaviruses^13^. To determine if antibodies against the S protein of hCoVs are boosted upon SARS-CoV-2 infection, we measured 229E, NL63, OC43, and SARS-CoV-2 S IgG antibody levels in sera collected longitudinally from 27 hospitalized COVID-19 patients. Serum IgG antibodies reactive to the S protein of the 229E and NL63 alphacoronaviruses did not change over 7 days of hospitalization (**Figure 3A-B**). Conversely, serum antibodies reactive to the S protein of OC43 and SARS-CoV-2 betacoronaviruses significantly increased over the course of hospitalization (**Figure 3A-B**). The magnitude of OC43 S antibody boost was not associated with outcome of disease (**Figure 3C**). Taken together, these data suggest that cross-reactive antibodies elicited by previous hCoV infections are not associated with protection from SARS-CoV-2 infections, but are boosted following infection with SARS-CoV-2.

**Figure 3.**
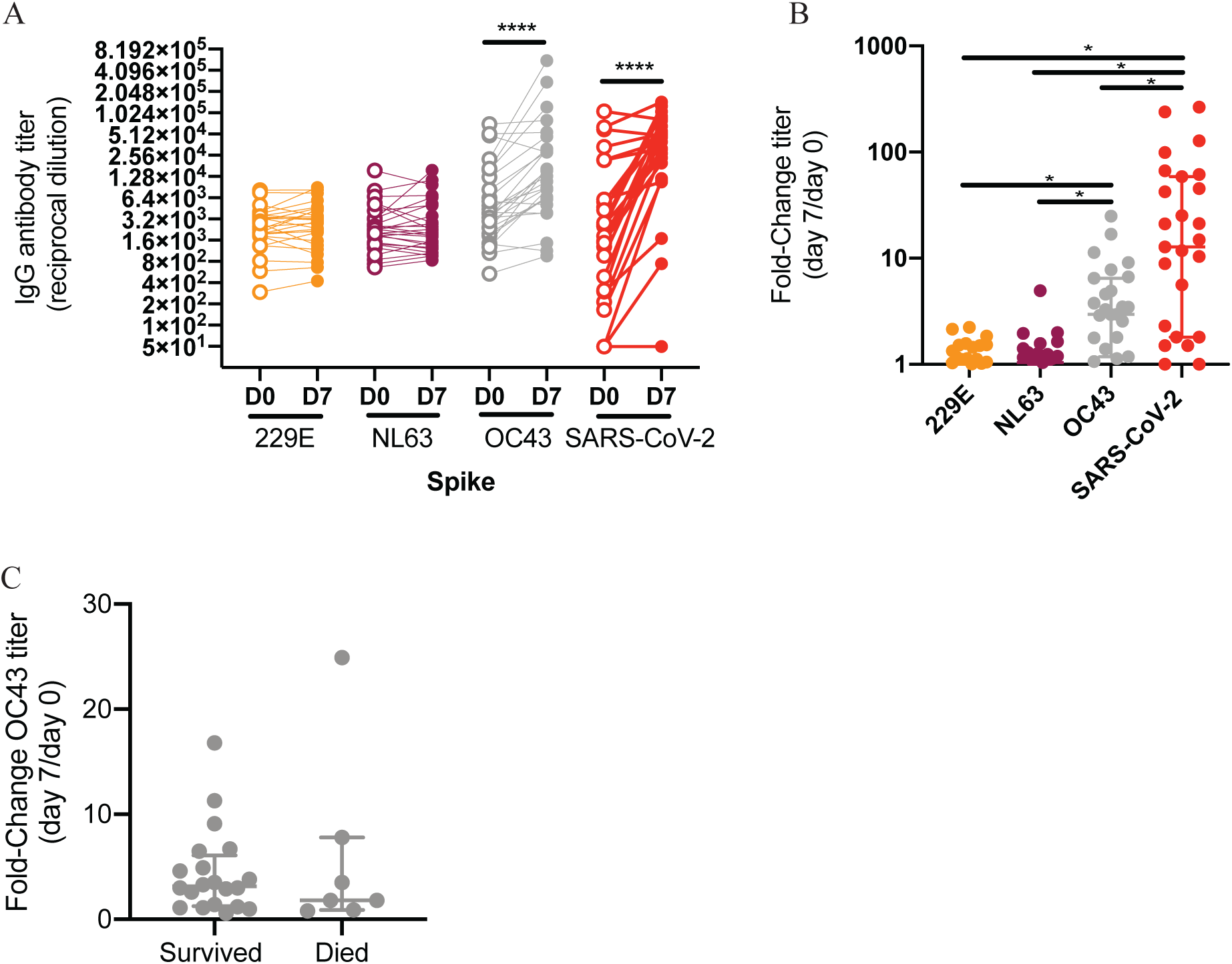
SARS-CoV-2 infections boost antibodies that react to OC43 S protein. We quantified antibody levels in serum collected from 27 individuals 0 and 7 days after hospitalization for COVID-19. ELISAs were completed to quantify levels of antibodies reactive to the S proteins of 229E, NL63, OC43 and SARS-CoV-2. (**A**) IgG titers and (**B**) titer fold change are shown. (**C**) Fold change in OC43 S-reactive antibodies was not associated with disease outcome. Paired t-test of log2 transformed antibody titers, ****p<0.0001. One-way ANOVA Tukey’s multiple comparisons fold-change in antibody titers, *p<0.04. Horizontal lines indicate the mean and error bars show standard deviation.

## DISCUSSION

Our study demonstrates that ∼23% of individuals possessed SARS-CoV-2 cross-reactive serum antibodies prior to the COVID-19 pandemic. Using samples collected in 2017, we found that pre-pandemic cross-reactive antibodies directed against the SARS-CoV-2 N protein were more prevalent compared to those directed against the SARS-CoV-2 S protein (18.6% seropositive versus 5.4% seropositive). We found that most individuals possessed pre-pandemic serum antibodies reactive to the S proteins of 229E, NL63, and OC43 (**Figure S2**); however, pre-pandemic samples with detectable levels of SARS-CoV-2 antibodies had higher levels of antibodies against the OC43 S protein (**Figure 1H**). Although our data suggest that prior infections with seasonal human betacoronaviruses (such as OC43) likely elicit antibodies that cross-react with SARS-CoV-2 proteins, in is unclear why only a subset of OC43 seropositive individuals possessed antibodies reactive to SARS-CoV-2 prior to the pandemic. Further studies will be needed to determine the temporal relationship between seasonal human betacoronavirus infections and the induction of SARS-CoV-2 cross-reactive antibodies. Further studies investigating the relationship of pre-pandemic antibodies against other betacoronaviruses, such as HKU1, with pre-pandemic SARS-CoV-2 cross-reactive antibodies are also needed.

We show that pre-pandemic SARS-CoV-2 cross-reactive antibodies are non-neutralizing and are not associated with reducing SARS-CoV-2 infections and hospitalizations. We compared serum from individuals who were and were not hospitalized after SARS-CoV-2 infections and found no differences in pre-pandemic antibody levels against SARS-CoV-2 and OC43 (**Figure 2**). We evaluated the need for respiratory support and admittance into the ICU as a proxy for COVID-19 severity (**Table S2**); however, larger cohorts including individuals with a large range of different clinically-defined disease severities will be required to determine if pre-pandemic levels of antibodies are associated with reducing some aspects of severe COVID-19. Additional studies need to be completed to determine if neutralizing antibodies elicited by SARS-CoV-2 infections protect against subsequent reinfections with SARS-CoV-2.

Further studies also need to be completed to determine how immune history affects de novo immune responses following SARS-CoV-2 infection. We find that individuals infected with SARS-CoV-2 produce antibodies reactive to both the SARS-CoV-2 S protein and OC43 S protein (**Figure 3**). In the case of influenza viruses, sequential infections with antigenically distinct strains can elicit antibodies against conserved epitopes between the strains and it is unclear if these cross-reactive antibodies inhibit de novo immune responses or affect disease severity^15^. Further studies are needed to precisely map the footprints of OC43 S-reactive antibodies elicited by SARS-CoV-2 infections. Additional studies need to be completed to determine if these antibodies help resolve infections or if they enhance disease in COVID-19 patients.

## STAR METHODS

### KEY RESOURCES TABLE

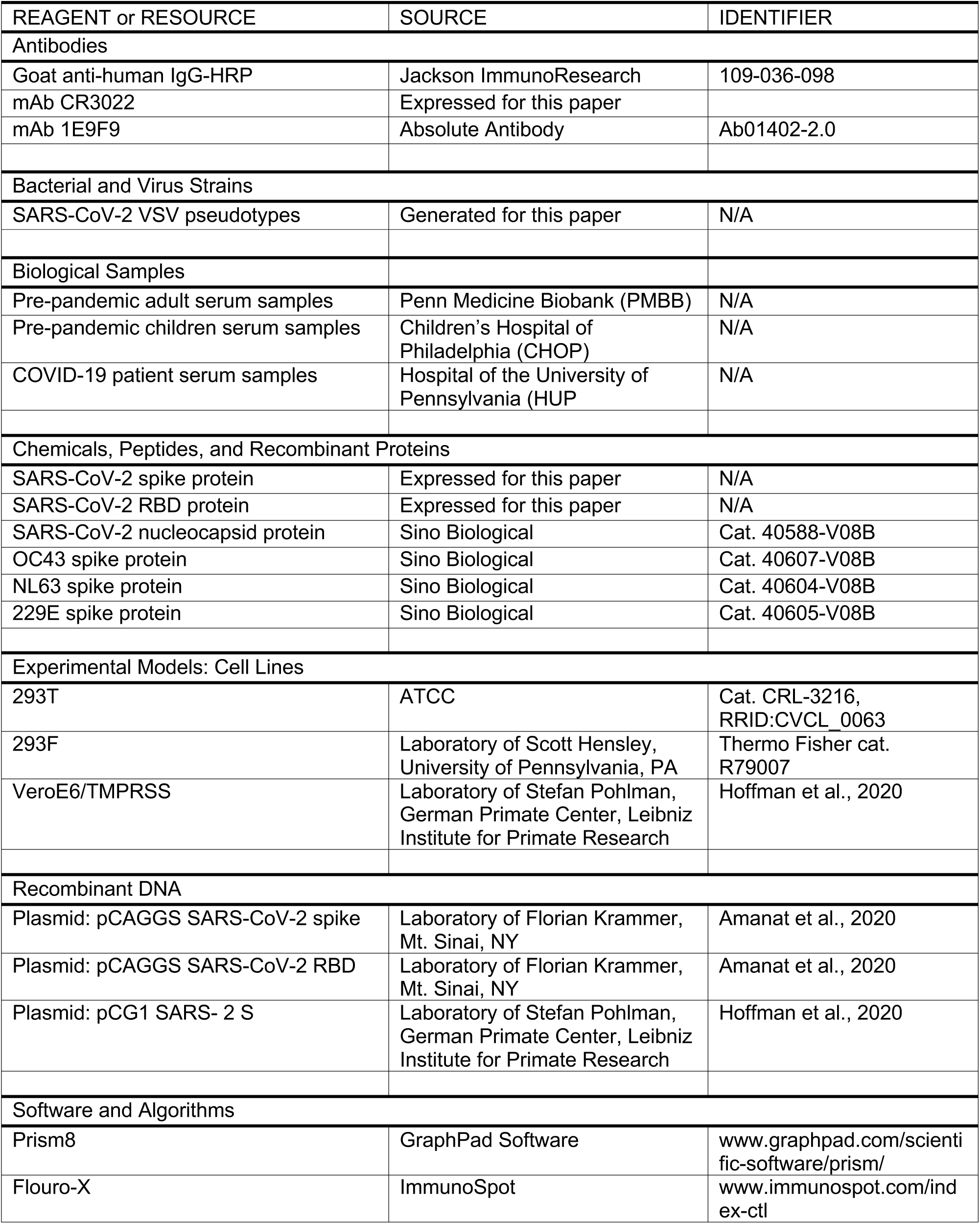

### RESOURCES AVAILABILITY

#### Lead Contact

Further information and requests for resources and reagents should be directed to and will be fulfilled by the Lead Contact, Scott E. Hensley.

#### Materials Availability

All unique reagents generated in this study will be available from the Lead Contact upon reasonable request.

#### Data and Code Availability

The published article includes all data generated or analyzed during this study.

## EXPERIMENTAL MODEL AND SUBJECT DETAILS

### Pre-pandemic Human Serum Samples

Serum samples shown in **Figure 1** were collected before the COVID-19 pandemic between May and August of 2017 from individuals at the Children’s Hospital of Philadelphia (CHOP; n=36, children age 0-18 years old) and through the Penn Medicine BioBank (n=168, adults ≥18 years old). Samples from CHOP were leftover de-identified blood samples collected for routine lead testing.

Serum samples shown in **Figure 2** were collected via the Penn Medicine BioBank prior to the pandemic (n=502, between August 2013 and March 2020). These samples were from adults who subsequently had a reverse transcription quantitative polymerase chain reaction (RT-qPCR) confirmed SARS-CoV-2 infection using nasopharyngeal swabs (cases, n=251), and those who had SARS-CoV-2 PCR negative results (controls, n=251). The RT-qPCR clinical testing results were acquired from Penn Medicine electronic health records and test results between March 2020 and August 2020 were included in the analysis. The Penn Medicine BioBank is an established repository that routinely collects blood products from donors visiting the University of Pennsylvania Healthcare system upon written informed consent. All studies were approved by the University of Pennsylvania Institutional Review Board.

### Human Samples Collected After SARS-CoV-2 Infection

Serum samples were obtained from recovered convalescent donors who had a history of PCR-confirmed SARS-CoV-2 infection (n=15). These samples were used in experiments shown in Figure 1. Additionally, plasma samples were collected from patients admitted to the Hospital at the University of Pennsylvania (HUP) with PCR-confirmed SARS-CoV-2 infections (n=27), as previously described^16^. Hospital inpatients were categorized for pneumonia severity using a WHO ordinal scale that was based on the level of oxygen support needed at day 0 and day 7. All samples were collected after obtaining informed consent and studies were approved by the University of Pennsylvania Institutional Review Board.

### Cell lines

293F cells were from Thermo fisher (Thermo Fisher cat. R79007). 293T cells were from ATCC (ATCC cat. CRL-3216, RRID:CVCL_0063).VeroE6/TMPRSS2 cells were a gift from Stefan Pohlman (German Primate Center, Leibniz Institute for Primate Research) as described previously^17^. All cell lines were cultured using manufacturer’s guidelines and used as described in Method Details below.

## METHOD DETAILS

### Quantification of serum antibody titers

Serum antibody titers against SARS-CoV-2 and other human coronavirus (hCoV) antigens were quantified by enzyme-linked immunosorbent assays (ELISA) as previously described^18^. Plasmids encoding the full-length SARS-CoV-2 spike (S) protein and the receptor binding domain of the S (S-RBD) were provided by Florian Krammer (Icahn School of Medicine at Mt. Sinai, New York City NY)^19^. SARS-CoV-2 S-RBD and the SARS-CoV-2 S proteins were purified from 293F transfected cells by Ni-NTA resin. SARS-CoV-2 nucleocapsid (N) protein, and full-length hCoV spike antigens (OC43, 229E, and NL63) were purchased (Sino Biological, Wayne PA; cat. 40588-V08B, 40607-V08B, 40604-V08B, and 40605-V08B, respectively) and reconstituted in Dulbecco’s phosphate buffered saline (DPBS). ELISA plates (Thermo Fisher Scientific: cat. 14-245-153) were coated overnight at 4°C with either 2 μg/mL SARS-CoV-2 antigen, 1.5 μg/mL hCOV antigen, or DPBS to control for background. Sera was heat-inactivated in a 56°C water bath for 1 hour prior to serial dilutions starting at 1:50 in dilution buffer (DPBS supplemented with 1% milk and 0.1% Tween-20). ELISA plates were blocked with 200μL of blocking buffer (DPBS supplemented with 3% milk and 0.1% Tween-20), washed 3 times with PBS plus 2% Tween (PBS-T), and 50μL of diluted sera was added. After 2 hours of incubation, ELISA plates were washed 3 times with PBS-T and bound antibodies were detected using a 1:5000 dilution of goat anti-human IgG conjugated to horseradish peroxidase (Jackson ImmunoResearch Laboratories, West Grove PA: cat. 109-036-098). ELISA plates were developed with the addition of 50 μL SureBlue 3, 3’, 5, 5’-tetramethylbenzidine substrate (SeraCare: material number 5120-0077) and the reactions were stopped by the addition of 25μL of 250mM hydrochloric acid after 5 minutes. Optical densities at 450nm wavelength were obtained on a SpectraMax 190 microplate reader (Molecular Devices, San Jose CA). Serum antibody titers were expressed as the reciprocal serum dilution at a set OD that was based off of a standard curve from the monoclonal antibody CR3022 (a gift from Ian Wilson, Scripps) starting at 0.5μg/mL (for S-RBD and S ELISAs) or serially diluted pooled serum (for SARS-CoV-2 N ELISAs and hCoV S ELISAs). Standard curves were included on every plate to control for plate-to-plate variation. Antibody titers for each sample were measured in at least two technical replicates performed on separate days.

### Generation of SARS-CoV-2 pseudotypes

SARS-CoV-2 pseudotypes were generated with a previously described vesicular stomatitis virus (VSV) pseudotype platform^20^. Briefly, pseudotyped VSV virions with SARS-CoV-2 Spike were produced through transfection of 293T with 35μg of pCG1 SARS-CoV-2 S delta18 expression plasmid encoding a codon optimized SARS-CoV-2 S gene with an 18-residue truncation in the cytoplasmic tail (kindly provided by Stefan Pohlmann)^17^. 30 hours post transfection, the SARS-CoV-2 spike expressing cells were infected for 2-4 hours with VSV-G pseudotyped VSVΔG-RFP at a multiplicity of infection (MOI) of ∼1-3. Then, the cells were washed twice with media to remove unbound virus. 28-30 hours after infection, the media containing the VSVΔG-RFP SARS-CoV-2 pseudotypes were harvested and clarified by centrifugation two times at 6000xg. SARS-CoV-2 pseudotypes were aliquoted and stored at −80°C until used for antibody neutralization analysis.

### Quantification of SARS-CoV-2 neutralizing antibody titers

Serum SARS-CoV-2 neutralizing antibodies were measured as previously described^20^. Vero E6 cells stably expressing TMPRSS2 were seeded in 100μl at 2.5×10^4^ cells/well in a 96 well collagen coated plate. The next day, heat inactivated serum samples were serially diluted 2-fold and mixed with 50-200 focus forming units/well of VSVΔG-RFP SARS-CoV-2 pseudotype virus and 600ng/ml of 1E9F9, a mouse anti-VSV Indiana G (Absolute Antibody, Oxford, UK: cat# Ab01402-2.0). The serum-virus mixture was incubated for 1 hour at 37⁰C before being plated on VeroE6 TMPRSS2 cells. 23-24 hours post infection, the cells were washed, fixed with 4% paraformaldehyde, and visualized on an S6 FluoroSpot Analyzer (CTL, Shaker Heights OH) and individual infected foci were enumerated. The focus reduction neutralization titer 50% (FRNT_50_) was measured as the greatest serum dilution at which focus count was reduced by at least 50% relative to control cells that were infected with pseudotype virus in the absence of human serum. FRNT_50_ titers for each sample were measured in at least two technical replicates performed on separate days.

## QUANTIFICATION AND STATISTICAL ANALYSIS

Statistical analyses were performed using Prism version 8 (GraphPad Software, San Diego CA). Reciprocal serum dilution antibody titers were log2 transformed for statistical analysis. ELISA antibody titers below the limit of detection (LOD; reciprocal titer <50) were set to a reciprocal titer of 25. Log2 transformed antibody titers were compared with unpaired t-tests and statistical significance was set to p-value <0.05. Linear regressions were also performed using log2 transform titers and untransformed data from the other variables. We compared antibody titers in pre-pandemic serum samples from individuals who did and did not have a subsequent PCR-confirmed SARS-CoV-2 infection. For these analyses we selected serum sample from individuals with RT-PCR negative results matching sex, age, and race for each SARS-CoV-2 PCR-confirmed case (RT-PCR positive) to define controls for our cohort. In instances we did not find matched controls, we randomly selected patients with RT-PCR negative test results. We also compared antibody titers in pre-pandemic serum samples among SARS-CoV-2 PCR-confirmed individuals in relationship to hospitalization or need for respiratory support due to COVID-19. Multivariate logistic regression was used to compare the antibody differences for these studies. All the models were adjusted by sex, age, race, and analyses were performed in R^21^. We compared Log2 transformed antibody titers in COVID-19 hospitalized patients at day 0 and day 7. We also compared the fold change in titer by day 7. We compared the fold change in OC43 titers between patients who survived and patients who died by day 28 of hospitalization.

## Data Availability

The article includes all data generated or analyzed during this study.

## ACKNOWLEDGEMENTS

EMA and TBM were supported by the NIH Training in Virology T32 Program through grant number T32-AI-007324. PH was supported by the NIH Emerging Infectious Diseases T32 Program T32-AI055400. PB was supported by a Peer Reviewed Medical Research Program award PR182551 and grants from the NIH (R21AI129531 and R21AI142638). This work was supported by institutional funds from the University of Pennsylvania and NIH HL137006 (NJM) and HL137915 (NJM). We thank J. Lurie, J. Embiid, J. Harris, and D. Blitzer for philanthropic support. We thank all members of the Wherry Lab and the Penn COVID-19 Sample Processing Unit (Zahidul Alam, Mary M. Addison, Katelyn T. Byrne, Aditi Chandra, Hélène C. Descamps, Yaroslav Kaminskiy, Jacob T. Hamilton, Julia Han Noll, Dalia K. Omran, Eric Perkey, Elizabeth M. Prager, Dana Pueschl, Jennifer B. Shah, Jake S. Shilan, Ashley N. Vanderbeck) for sample procurement, processing, and logistics. We thank the staff of the PMBB. We thank F. Krammer (Mt. Sinai) for sending us the SARS-CoV-2 spike RBD expression plasmids.

## FIGURES

**Figure S1.**
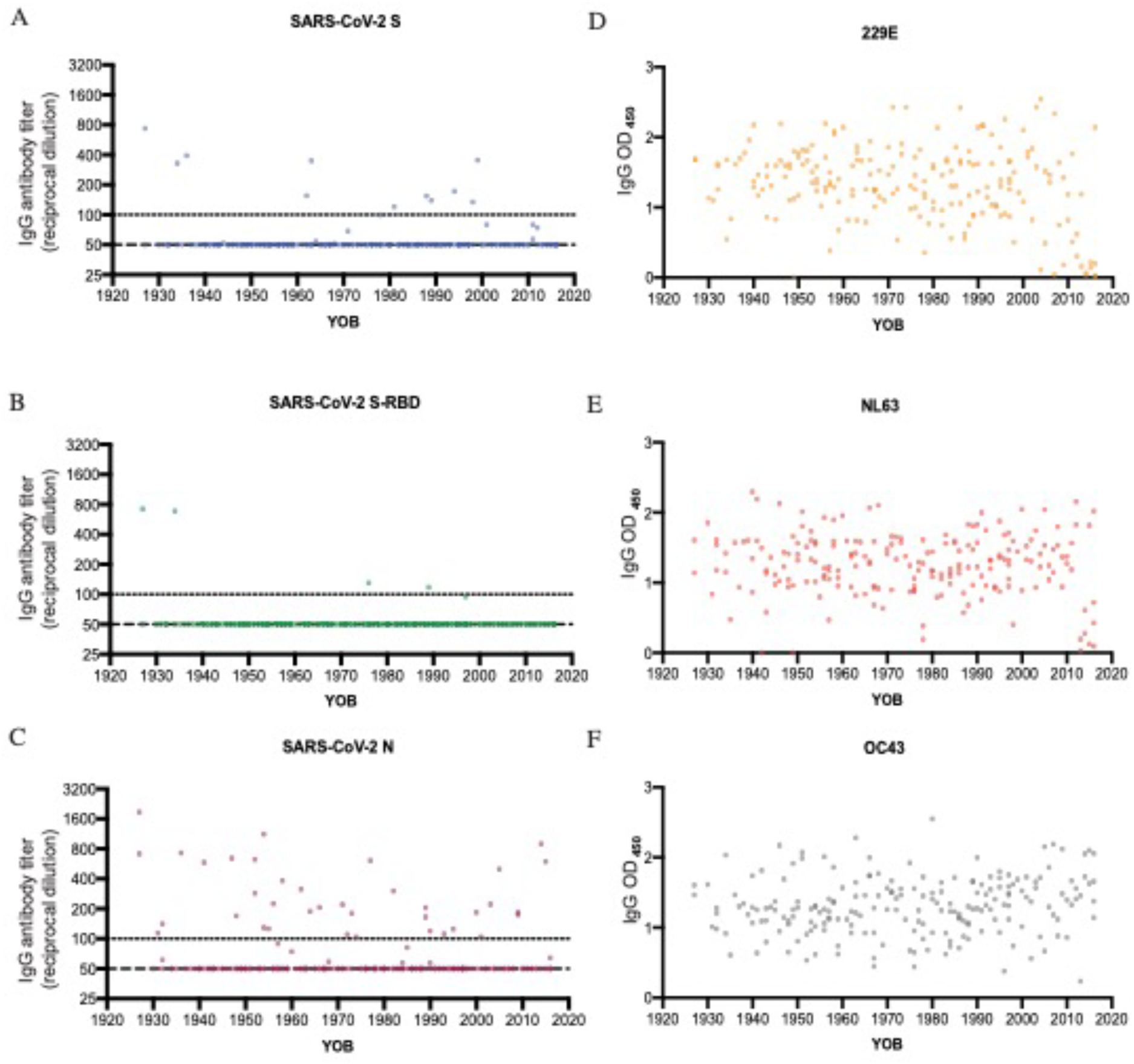
There are no obvious age-related differences in pre-pandemic SARS-CoV-2 and hCoV reactive antibodies. ELISAs were completed to measure levels of serum antibodies binding to the SARS-CoV-2 full-length spike (S) protein (**A**), SARS-CoV-2 receptor binding domain (S-RBD) of S (**B**), SARS-CoV-2 nucleocapsid (N) protein (**C**), 229E S protein (**D**), NL63 S protein (**E**), and OC43 S protein (**F**). Serum samples collected from 204 individuals in the summer of 2017 were tested. Reciprocal titer from serially-diluted serum samples (**A-C**) and optical densities at 450nm wavelength (OD_450_) of 1:500 dilution of serum (**D-F**) are shown. Dashed line denotes lower limit of detection (LOD=50), dotted line represents a threshold set 2-fold above LOD (>100).

## Supplementary Tables

**Table S1:**
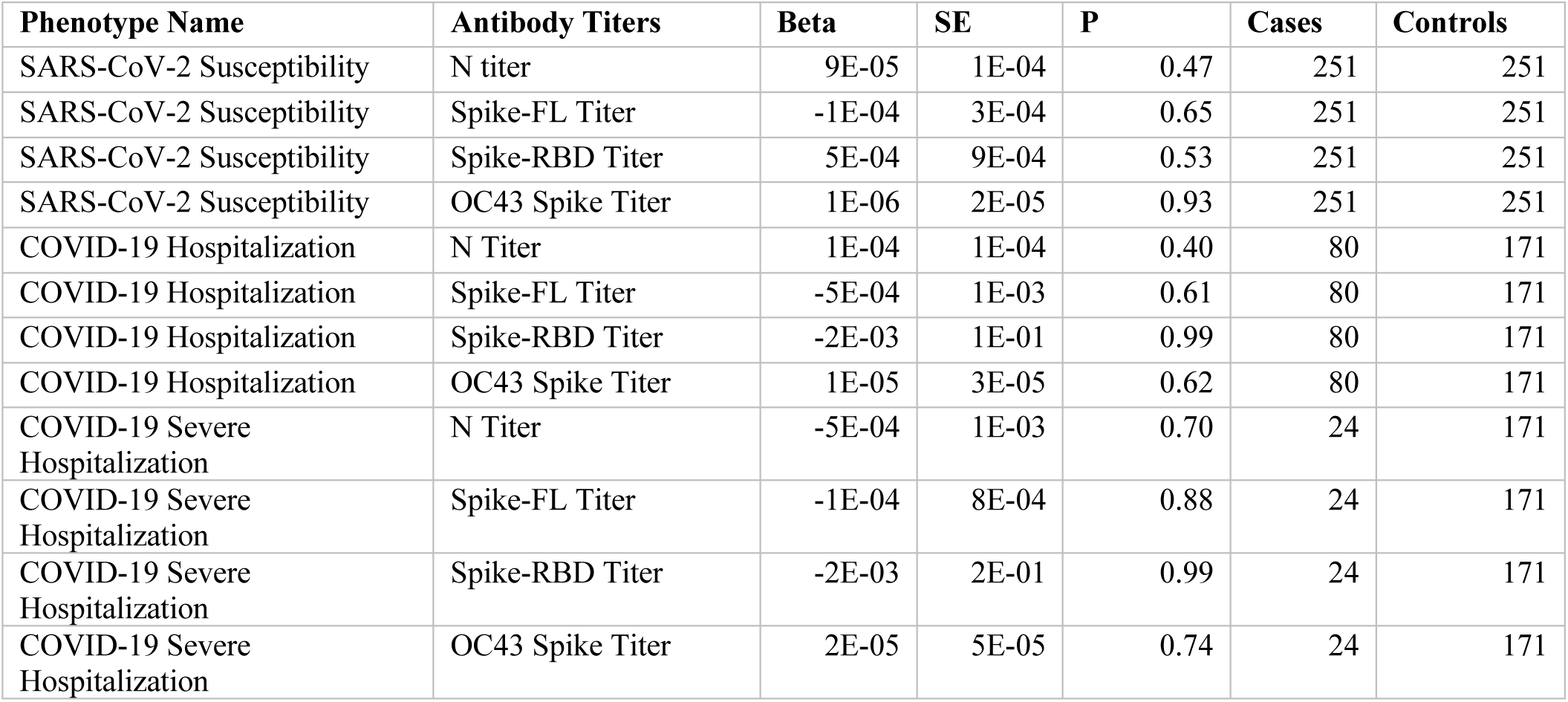
Comparison between antibody titers and COVID-19 phenotypes.

| Phenotype Name | Antibody Titers | Beta | SE | P | Cases | Controls |
| --- | --- | --- | --- | --- | --- | --- |
| SARS-CoV-2 Susceptibility | N titer | 9E-05 | 1E-04 | 0.47 | 251 | 251 |
| SARS-CoV-2 Susceptibility | Spike-FL Titer | -1E-04 | 3E-04 | 0.65 | 251 | 251 |
| SARS-CoV-2 Susceptibility | Spike-RBD Titer | 5E-04 | 9E-04 | 0.53 | 251 | 251 |
| SARS-CoV-2 Susceptibility | OC43 Spike Titer | 1E-06 | 2E-05 | 0.93 | 251 | 251 |
| COVID-19 Hospitalization | N Titer | 1E-04 | 1E-04 | 0.40 | 80 | 171 |
| COVID-19 Hospitalization | Spike-FL Titer | -5E-04 | 1E-03 | 0.61 | 80 | 171 |
| COVID-19 Hospitalization | Spike-RBD Titer | -2E-03 | 1E-01 | 0.99 | 80 | 171 |
| COVID-19 Hospitalization | OC43 Spike Titer | 1E-05 | 3E-05 | 0.62 | 80 | 171 |
| COVID-19 Severe Hospitalization | N Titer | -5E-04 | 1E-03 | 0.70 | 24 | 171 |
| COVID-19 Severe Hospitalization | Spike-FL Titer | -1E-04 | 8E-04 | 0.88 | 24 | 171 |
| COVID-19 Severe Hospitalization | Spike-RBD Titer | -2E-03 | 2E-01 | 0.99 | 24 | 171 |
| COVID-19 Severe Hospitalization | OC43 Spike Titer | 2E-05 | 5E-05 | 0.74 | 24 | 171 |

**Table S2:** Phenotype definitions related to Table S1.

| Phenotype Name | Case Definition | Control Definition | Case | Controls |
| --- | --- | --- | --- | --- |
| SARS-CoV-2 Susceptibility | RT-PCR confirmed *positive* test for SARS-CoV2 infection | RT-PCR confirmed *negative* test for SARS-CoV2 infection | 251 | 251 |
| COVID-19 Hospitalization | RT-PCR confirmed positive test for SARS-CoV2 infection and *hospitalized* due to COVID-19 | RT-PCR confirmed positive test for SARS-CoV2 infection and *not hospitalized* due to COVID-19 | 80 | 171 |
| COVID-19 Severe Hospitalization | RT-PCR confirmed positive test for SARS-CoV2 infection and *required respiratory support or had ICU stay* due to COVID-19 | RT-PCR confirmed positive test for SARS-CoV2 infection and *not hospitalized* due to COVID-19 | 24 | 171 |

